# Antibody response durability following three-dose COVID-19 vaccination in people with HIV receiving suppressive ART

**DOI:** 10.1101/2022.11.03.22281912

**Authors:** Hope R. Lapointe, Francis Mwimanzi, Peter K. Cheung, Yurou Sang, Fatima Yaseen, Sarah Speckmaier, Evan Barad, Nadia Moran-Garcia, Sneha Datwani, Maggie C. Duncan, Rebecca Kalikawe, Siobhan Ennis, Landon Young, Bruce Ganase, F. Harrison Omondi, Gisele Umviligihozo, Winnie Dong, Junine Toy, Paul Sereda, Laura Burns, Cecilia T. Costiniuk, Curtis Cooper, Aslam H. Anis, Victor Leung, Daniel Holmes, Mari L. DeMarco, Janet Simons, Malcolm Hedgcock, Natalie Prystajecky, Christopher F. Lowe, Marc G. Romney, Rolando Barrios, Silvia Guillemi, Chanson J. Brumme, Julio S.G. Montaner, Mark Hull, Marianne Harris, Masahiro Niikura, Mark A. Brockman, Zabrina L. Brumme

**Affiliations:** British Columbia Centre for Excellence in HIV/AIDS, Vancouver, Canada; Faculty of Health Sciences, Simon Fraser University, Burnaby, Canada; Department of Molecular Biology and Biochemistry, Simon Fraser University, Burnaby, Canada; Division of Medical Microbiology and Virology, St. Paul’s Hospital, Vancouver, Canada; AIDS Research Program, St. Paul’s Hospital, Vancouver, Canada; Department of Pathology and Laboratory Medicine, Providence Health Care, Vancouver, Canada; Division of Infectious Diseases and Chronic Viral Illness Service, McGill University Health Centre and Research Institute of the McGill University Health Centre, Montreal, Quebec, Canada; Department of Medicine, University of Ottawa, Ottawa, Canada; Ottawa Hospital Research Institute, Ottawa, Canada; School of Population and Public Health, University of British Columbia, Vancouver, Canada; CIHR Canadian HIV Trials Network, University of British Columbia, Vancouver, Canada; Centre for Health Evaluation and Outcome Sciences, Vancouver, Canada; Department of Pathology and Laboratory Medicine, University of British Columbia, Vancouver, Canada; Spectrum Health, Vancouver, Canada; British Columbia Centre for Disease Control Public Health Laboratory, Vancouver, Canada; Department of Family Practice, Faculty of Medicine, University of British Columbia, Canada; Department of Medicine, University of British Columbia, Vancouver, Canada

**Author notes:** **Corresponding Author Contact Information:** Zabrina L. Brumme, Ph.D., Professor, Faculty of Health Sciences, Simon Fraser University, 8888 University Drive, Burnaby, BC, Canada, V5A 1S6. denote equal contribution (Hope R. Lapointe, Francis Mwimanzi, Peter K. Cheung, Mark A. Brockman, Zabrina L. Brumme).

**Keywords:** HIV, COVID-19, vaccine, Omicron BA.1, Omicron BA.5, vaccines, humoral immunity, hybrid immunity, antibody, viral neutralization, third dose

## Abstract

**Background:** Limited data exist regarding longer-term antibody responses following three-dose COVID-19 vaccination, and the impact of a first SARS-CoV-2 infection during this time, in people living with HIV (PLWH) receiving suppressive antiretroviral therapy (ART). We quantified wild-type-(WT), Omicron BA.1- and Omicron BA.5-specific responses up to six months post-third dose in 64 PLWH and 117 controls who remained COVID-19-naive or experienced their first SARS-CoV-2 infection during this time.

**Design:** Longitudinal observational cohort.

**Methods:** We quantified WT- and Omicron-specific Anti-Spike receptor-binding domain IgG concentrations, ACE2 displacement activities and live virus neutralization at one, three and six months post-third vaccine dose.

**Results:** Third doses boosted all antibody measures above two-dose levels, but BA.1-specific responses remained significantly lower than WT-specific ones, with BA.5-specific responses lower still. Serum IgG concentrations declined at similar rates in COVID-19-naive PLWH and controls post-third dose (median WT- and BA.1-specific half-lives were between 66-74 days for both groups). Antibody function also declined significantly yet comparably between groups: six months post-third dose, BA.1-specific neutralization was undetectable in >80% of COVID-19 naive PLWH and >90% of controls. Breakthrough SARS-CoV-2 infection boosted antibody concentrations and function significantly above vaccine-induced levels in both PLWH and controls, though BA.5-specific neutralization remained significantly poorer than BA.1 even post-breakthrough.

**Conclusions:** Following three-dose COVID-19 vaccination, antibody response durability in PLWH receiving ART is comparable to controls. PLWH also mounted strong responses to breakthrough infection. Due to temporal response declines however, COVID-19-naive individuals, regardless of HIV status, would benefit from a fourth dose within 6 months of their third.

## INTRODUCTION

In British Columbia (BC), Canada, third doses of COVID-19 monovalent mRNA vaccines were introduced in November 2021, initially to individuals at risk of severe COVID-19 outcomes, including some people living with HIV (PLWH). Whether offered as part of a primary vaccine series or a “booster”, third doses help to maintain systemic immunity and enhance protection against infection by viral variants [1-4]. Despite being effective at preventing severe disease due to SARS-CoV-2 however, third doses provide limited protection against transmission of Omicron subvariants, including BA.1 and BA.5 [5-10], which were estimated to infect more than 60% of Canadians by August 2022, despite approximately 50% uptake of third doses [11-13].

Longitudinal monitoring of immune responses post-third dose in PLWH is critical to inform the timing of future immunizations in this group. Though some data are available on initial immunogenicity to third COVID-19 vaccine doses in PLWH [14, 15], no studies to our knowledge have assessed the longer-term durability of responses in this population. Furthermore, despite the high incidence of first-time SARS-CoV-2 infections after three vaccine doses, no studies to our knowledge have examined the impact of such infections on responses in PLWH. Here, we extend prior observations from our cohort [14, 16] by quantifying wild-type-(WT), Omicron BA.1- and BA.5-specific responses up to six months post-third vaccine dose in 64 PLWH and 117 controls who either remained COVID-19-naive or experienced their first (presumably Omicron [17]) SARS-CoV-2 infection, during this period.

## METHODS

### Participants

Our cohort was described previously [14]. The present study included 64 PLWH and 117 controls who remained COVID-19-naive until at least one month post-third vaccine dose. Breakthrough SARS-CoV-2 infections were identified through self-reported PCR and/or rapid-antigen test results and the presence of serum antibodies against Nucleocapsid (N) using the Elecsys Anti-SARS-CoV-2 assay (Roche Diagnostics).

### Ethics approval

This study was approved by the University of British Columbia/Providence Health Care and Simon Fraser University Research Ethics Boards. All participants provided written informed consent.

### Antibody assays

Assays were performed as previously described [14, 18]. IgG-binding antibodies in serum were measured against the SARS-CoV-2 Spike Receptor Binding Domain (RBD) using the V-plex SARS-CoV-2 (IgG) ELISA kit (Panel 22; Meso Scale Diagnostics), which features WT and Omicron-BA.1 RBD antigens, on a Meso QuickPlex SQ120 instrument. Serum was diluted 1:10000 and reported in World Health Organization (WHO) International Standard Binding Antibody Units (BAU)/mL using the manufacturer-supplied conversions. Surrogate virus neutralization activity [19] in serum was measured by competition ELISA using the same kit (Panel 22; V-plex SARS-CoV-2 [ACE2]) to measure blockade of the RBD-ACE2 receptor interaction. Sera were diluted 1:40 and results reported as % ACE2 displacement. Virus neutralizing activity in plasma was assessed using live WT (USA-WA1/2020; BEI Resources), and two local isolates identified as Omicron BA.1 (GISAID Accession# EPI_ISL_9805779) and Omicron BA.5 (GISAID Accession# EPI_ISL_15226696) on VeroE6-TMPRSS2 (JCRB-1819) target cells [18]. Virus stocks were diluted to 50 TCID_50_/200 µl in the presence of serial 2-fold plasma dilutions (1/20 to 1/2560) and added to target cells in triplicate. Viral cytopathic effects (CPE) were recorded three days post-infection. Neutralization was reported as the highest reciprocal dilution able to prevent CPE in all three wells. Partial or no neutralization at 1/20 dilution was considered below the limit of quantification (BLOQ) and coded as a reciprocal dilution of 10.

### Statistical analyses

Continuous variables were compared using the Mann-Whitney U-test (unpaired data) or Wilcoxon test (paired data). Relationships between continuous variables were assessed using Spearman’s correlation. In participants who remained COVID-19 naive, multiple linear regression was used to investigate the relationship between HIV infection and vaccine-induced immune measures using a confounder model that adjusted for variables that could influence vaccine responses or that differed in prevalence between groups. For Omicron-specific neutralization at 6 months post-third dose, multiple logistic regression was used due to the high proportion of results BLOQ. Included variables were: HIV infection (controls as reference group), age (per year), sex at birth (female as reference), ethnicity (non-white as reference), number of chronic conditions (per additional), dual ChAdOx1 as the initial regimen (mRNA or mixed [ChAdOx1/mRNA] regimen as the combined reference group) [14, 16], third COVID-19 mRNA dose brand (BNT162b2 as reference) and the interval between second and third doses (per day). Plasma neutralization models also corrected for anticoagulant (ACD as reference). All tests were two-tailed, with p<0.05 considered statistically significant. Analyses were conducted using Prism v9.2.0 (GraphPad).

## RESULTS

### Participant characteristics

Characteristics of the 64 PLWH and 117 controls, all of whom remained COVID-19-naive until at least one month post-third dose, are shown in **Table 1**. All PLWH had suppressed plasma HIV viremia on ART, median CD4+ T-cell counts of 645 (Interquartile Range [IQR] 473-958) cells/mm^3^, and median nadir CD4+ T-cell counts of 225 (IQR 95-485) cells/mm^3^, at enrolment. PLWH were a median of 57 (IQR 42-65) years old and 90% male; controls were a median 47 (IQR 35-72) years old and 73% female. PLWH had a higher proportion of white ethnicity (72%, compared to 55% in controls) and more chronic health conditions (median 1 [IQR 1-3] compared to 0 [IQR 0-1] in controls). More PLWH (10%) than controls (<1%) received two doses of the recombinant viral vector ChAdOx1 vaccine as their initial immunization series. All third doses were monovalent mRNA vaccines, either BNT162b2 (30mcg) or mRNA-1273 (50 or 100mcg). Most PLWH (69%) and controls (62%) received mRNA-1273, where, per local guidelines, all adults aged ≥ 70 years were eligible for a 100mcg mRNA-1273 dose, as were PLWH who met one or more of the following criteria: age ≥ 65 years, prior AIDS-defining illness, prior CD4 count <200 cells/mm^3^, prior CD4 fraction ≤ 15%, any plasma HIV load >50 copies/mL in 2021, or perinatally-acquired HIV [20]. Third vaccine doses were administered approximately 6.5 months after the second dose.

**Table 1:** Participant characteristics.

| **Characteristic<sup>a</sup>** | **PLWH**<br>**(n=64)** | **Controls**<br>**(n=117)** |
| --- | --- | --- |
| **HIV-related variables** |  |  |
| Receiving antiretroviral therapy, n (%) | 64 (100%) | - |
| Recent plasma viral load, copies HIV RNA/mL, median [IQR] | <50 [<50 - <50] | - |
| Recent CD4+ T-cell count in cells/mm<sup>3</sup>, median [IQR] | 645 [473-958] | - |
| Nadir CD4+ T-cell count in cells/mm<sup>3</sup>, median [IQR] | 225 [95-485] | - |
| **Sociodemographic and health variables** |  |  |
| Age in years, median [IQR] | 57 [42-65] | 47 [35-72] |
| Female sex at birth, n (%) | 6 (9.4%) | 85 (73%) |
| white Ethnicity, n (%) | 46 (72%) | 64 (55%) |
| Number of chronic conditions, median [IQR]<sup>b</sup> | 1 [1-3] | 0 [0-1] |
| **Vaccine details** |  |  |
| Initial regimen |  |  |
| mRNA only | 67 (81.2%) | 114 (97.4%) |
| ChAdOx1 only | 6 (9.4%) | 1 (0.9%) |
| ChAdOx1/mRNA | 6 (9.4%) | 2 (1.7%) |
| Third dose |  |  |
| BNT162b2 | 20 (31.2%) | 45 (38.5%) |
| mRNA-1273 | 44 (68.8%) | 72 (61.5%) |
| Days between second and third doses, median [IQR] | 191 [182-240] | 198 [171-218] |
| **Specimen collection** |  |  |
| 1 month after third dose, n (%) | 63 (98%) | 116 (99%) |
| 3 months after third dose, n (%) | 58 (91%) | 116 (99%) |
| 6 months after third dose, n (%) | 55 (86%) | 110 (94%) |
| **Post-3rd dose SARS-CoV-2 infections, n (%)** | **24 (37.5%)** | **45 (38.5%)** |

<sup>a</sup>Sociodemographic, health and vaccine data were collected by self-report and confirmed through medical records where available.
<sup>b</sup>Chronic conditions were defined as hypertension, diabetes, asthma, obesity, chronic diseases of lung, liver, kidney, heart or blood, cancer, and immunosuppression due to chronic conditions or medication.

A total of 24 (38%) PLWH and 45 (39%) controls experienced their first SARS-CoV-2 infection between one and six months post-third dose (one control experienced two infections during this period [21]). Based on the 56 (81%) participants for whom infection timing was available, infections occurred a median 105 (IQR 76-137) days post-third dose, or a median date of 10-Apr-2022 (IQR 28-Feb–04-May), with no temporal differences between PLWH and controls (p=0.4). Though SARS-CoV-2 variant information is unavailable for individual infections, most were likely Omicron BA.1 or BA.2 based on local epidemiology at the time [17].

### Longitudinal binding IgG responses following three-dose vaccination

As reported previously, WT-specific serum IgG concentrations were comparable in COVID-19-naive PLWH and controls one month post-third dose (and also not significantly different post-second dose after adjustment for sociodemographic, health and vaccine-related variables [14, 16]) (**Figure 1A**). WT-specific IgG concentrations one month post-third dose were 3.70 (IQR 3.47-3.94) log_10_ BAU/mL in COVID-19-naive PLWH and 3.67 (IQR 3.50-3.86) log_10_ BAU/mL in controls, respectively (p=0.5). Following this, WT-specific IgG responses declined comparably in COVID-19-naive PLWH and controls: at three months, WT-specific IgG responses in PLWH and controls declined to a median 3.48 (IQR 3.12-3.75) and 3.40 (IQR 3.21-3.61) log_10_ BAU/mL, respectively (p=0.5), while by six months, responses had declined to a median 2.96 (IQR 2.62-3.38) and 3.06 (IQR 2.82-3.24) log_10_ BAU/mL in PLWH and controls, respectively (p=0.4) (**Figure 1A**; also see Figure **1B**). Indeed, by six months post-third dose, WT-specific IgG concentrations in COVID-19-naive PLWH had declined to levels comparable to those initially elicited by two vaccine doses (p=0.16), whereas those in controls had declined to significantly lower than post-second dose levels (p<0.0001) (**Figure 1A**).

**Figure 1.**
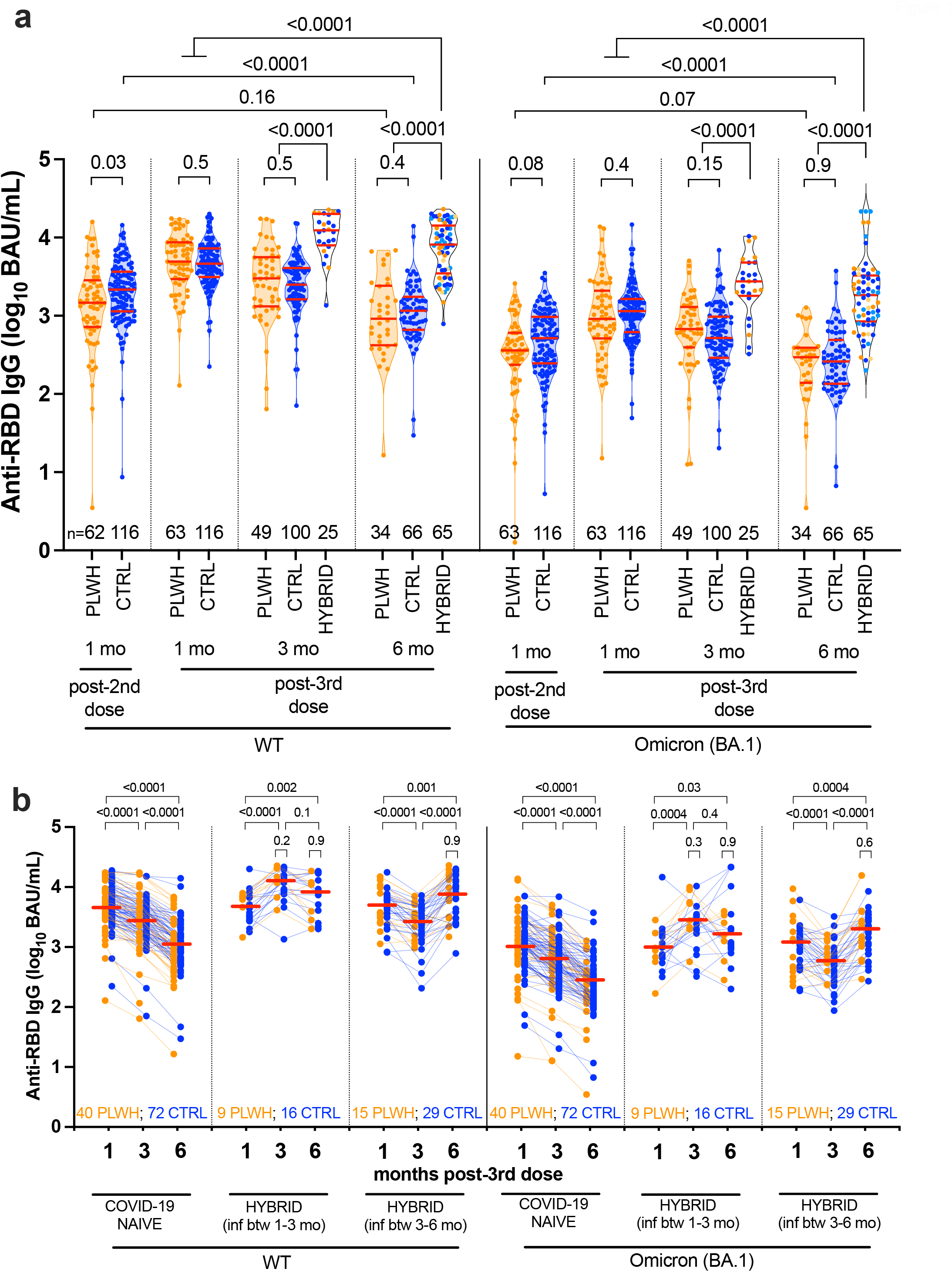
WT- and Omicron BA.1-specific anti-RBD IgG concentrations following three-dose COVID-19 vaccination. *Panel A:* Longitudinal serum anti-RBD IgG concentrations specific to WT (left side) and Omicron BA.1 (right side) in COVID-19-naїve people living with HIV (PLWH; orange circles) and controls (CTRL; blue circles). Any participant who experienced a SARS-CoV-2 breakthrough infection between 1-3 or 3-6 months post-third dose was reclassified into the “hybrid” group at their following study visit, with PLWH in orange and CTRL in blue circles. At six months post-third dose, the darker-colored symbols in the hybrid group denote recent infections that occurred between 3-6 months, while the lighter-colored symbols denote the infections that had previously occurred between 1-3 months. Red bars indicate median and IQR. Comparisons between independent groups were performed using the Mann-Whitney U-test; longitudinal paired comparisons were performed using the Wilcoxon matched pairs test. P-values are not corrected for multiple comparisons. *Panel B:* Same data as panel A, but where serum anti-RBD IgG concentrations are plotted longitudinally by participant (PLWH in orange, CTRL in blue). Participants are stratified into three groups: those who remained COVID-19-naїve throughout the study, those who experienced a SARS-CoV-2 breakthrough infection between 1-3 months post-third dose, those who experienced a SARS-CoV-2 breakthrough infection between 3-6 months post-third dose. Horizontal red lines denote the overall median response at each time point, where PLWH and CTRL are treated as a combined group. P-values above larger brackets compare responses between time points using the Wilcoxon matched pairs test, where PLWH and CTRL are treated as a combined group. P-values above small brackets compare responses between PLWH and CTRL at each time point post-SARS-CoV-2 infection using the Mann-Whitney U test. P-values are not corrected for multiple comparisons.

Similarly, BA.1-specific IgG responses were comparable in COVID-19-naive PLWH and controls at all post-third dose time points (all comparisons p≥0.15; **Figure 1A**). Nevertheless, BA.1-specific responses were significantly lower than WT-specific responses in all groups at all time points (all within-group comparisons of WT- and BA.1-specific responses were p<0.0001; not shown). One month post-third dose for example, BA.1-specific IgG concentrations were 2.96 (IQR 2.71-3.32) log_10_ BAU/mL in PLWH, which was 0.74 log_10_ BAU/mL lower than WT-specific concentrations at this time. By six months BA.1-specific IgG concentrations had declined to 2.47 (IQR 2.14-2.59) log_10_ BAU/mL in PLWH, which was 0.49 log_10_ BAU/mL lower than WT-specific concentrations at this time.

We next performed multivariable analyses adjusting for sociodemographic, health and vaccine-related variables to identify variables associated with WT- and BA.1-specific IgG concentrations at six months post-third dose in the COVID-19-naive subgroup. These analyses revealed that a higher number of chronic health conditions - but not HIV infection - was associated with poorer IgG responses at this time (p=0.028 for WT-specific responses; p= 0.016 for BA.1-specific responses), as was male sex (p=0.012 for BA.1-specific responses) (**Supplementary Table 1**). In fact, adjusted IgG concentrations were slightly *higher* in PLWH compared to controls. Moreover, receipt of an mRNA-1273 third dose (rather than BNT162b2) was associated with stronger WT-specific IgG responses (p=0.029), though not BA.1-specific responses (p=0.13). Among PLWH, we also observed no significant relationship between most recent or nadir CD4+ T-cell count and either WT- or BA.1-specific IgG concentrations at this time (p≥0.3; **Supplementary Figure 1**).

We estimated the half-lives of WT-specific IgG following three-dose vaccination to be a median 66 (IQR 47-89) days in COVID-19-naive PLWH compared to 72 (IQR 54-96) days in controls, a difference that was not statistically significant (p=0.2; **Supplementary Figure 2**). Estimated BA.1-specific IgG half-lives were also comparable, at a median 71 (IQR 49-104) days for PLWH compared to 74 (IQR 59-90) days in controls (p=0.8). Multivariable analyses confirmed that HIV infection was not associated with WT- or BA.1-specific IgG half-lives post-third dose (**Supplementary Table 2**). Among COVID-19-naive PLWH, we initially observed a weak inverse correlation between recent CD4+ T-cell count and IgG half-life, but this was not significant after excluding an outlier with a long (>200 day) half-life (**Supplementary Figure 1)**.

By contrast, the nearly 40% of PLWH and controls who experienced their first SARS-CoV-2 infection between one and six months post-third dose exhibited markedly higher WT- and BA.1-specific IgG concentrations than their COVID-19-naive counterparts at all post-infection time points (all comparisons p<0.0001 in **Figure 1A**; also see **Figure 1B**). In fact, at six months post-third dose, IgG responses in this “hybrid immunity” group were significantly higher than those initially induced by vaccination alone, *e*.*g*. BA.1-specific IgG concentrations were a median 3.26 log_10_ BAU/mL (IQR 2.93-3.52), which was 0.27 log_10_ BAU/mL higher than at one month post-third dose (p<0.0001). Importantly, “hybrid” IgG response magnitude was comparable between PLWH and controls (all p≥0.2; **Figure 1B**). Notably, while most participants experienced a marked boost in antibody levels following SARS-CoV-2 infection, IgG responses in a minority of PLWH and controls remained constant or even declined post-infection (**Figure 1B)**.

### Longitudinal ACE2 displacement activity following three-dose vaccination

One month post-third dose, WT-specific ACE2 displacement activity in COVID-19-naive PLWH was a median 99.6% (IQR 98.7-99.8%) compared to 99.1% (IQR 97.0-99.6%) in controls (univariable p=0.002; **Figure 2A**), though this did not remain significant after multivariable adjustment (not shown). Following this, WT-specific ACE2 displacement activities declined similarly in both COVID-19-naive groups: at three months, activities had decreased to a median 97.9% (IQR 88.8-99.6) in PLWH versus 98.8% (IQR 94.5-99.5) in controls (p=0.8), while by six months, activities had decreased to a median 87.0% (IQR 67.5-98.1) in PLWH versus 93.6% (IQR 81.5-97.8) in controls (p=0.3), levels that were comparable or lower than after two-dose vaccination (all p<0.1) (**Figures 2A** and **2B**). BA.1-specific responses remained significantly lower than WT-specific responses at all time points (all within-group comparisons for WT- and BA.1-specific responses p<0.0001; not shown), but these responses also declined similarly in COVID-19-naive PLWH and controls (**Figure 2A**). Between one and six months post-third dose for example, BA.1-specific ACE2 displacement activities decreased from a median 58.5% (IQR 33.2-78.2%) in PLWH and 63.4% (IQR 44.1-77.9%) in controls (p=0.4), to 30.8% (IQR 23.0-54.1) in PLWH and 25.9% (IQR 12.1-52.2) in controls (p=0.3), where the latter values were significantly lower than those observed after two-dose vaccination (all comparisons p≤0.02).

**Figure 2.**
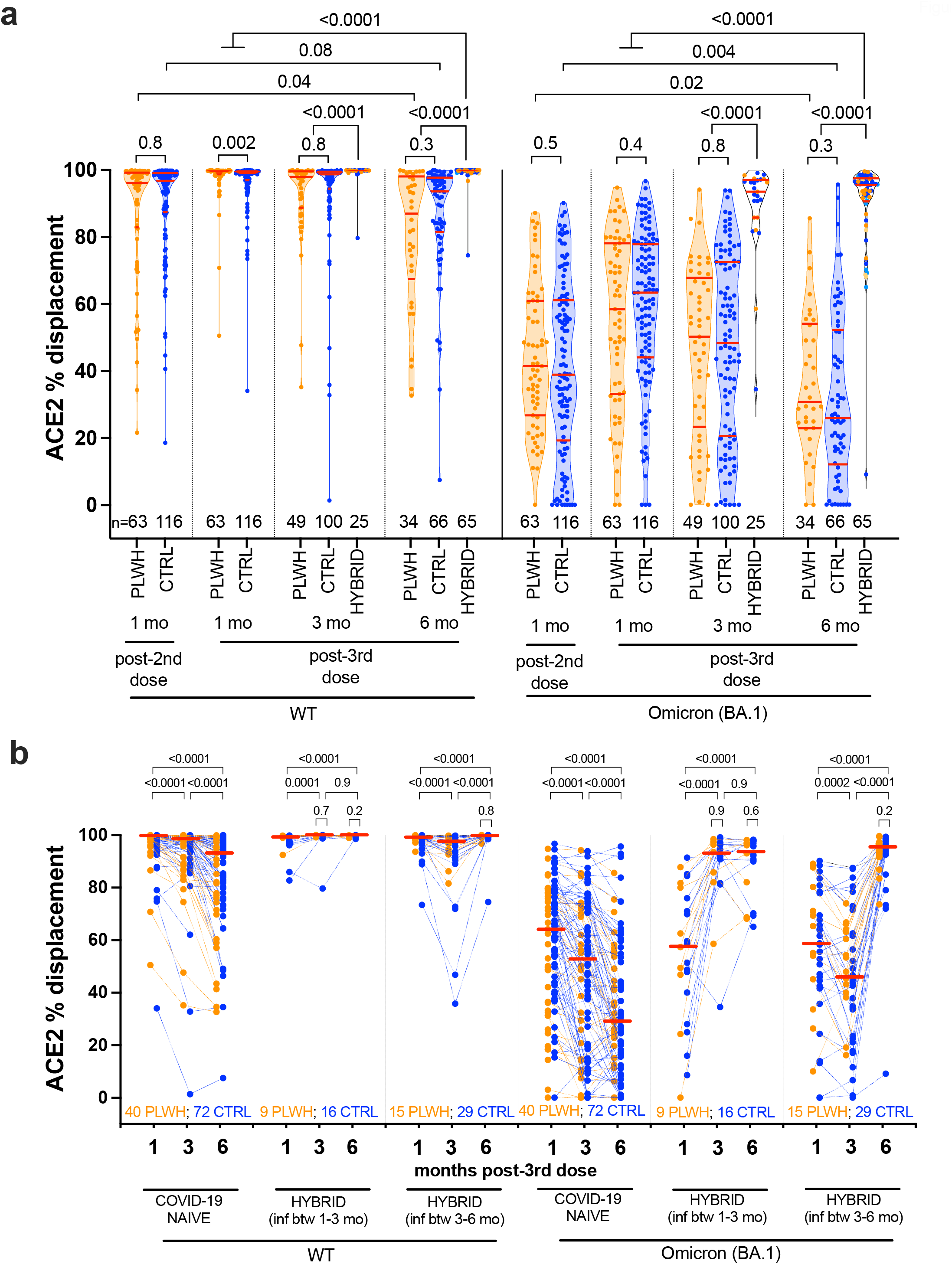
WT- and Omicron BA.1-specific ACE2 displacement function following three-dose COVID-19 vaccination. *Panel A:* Same as Figure 1A, but for ACE2 displacement activity in serum, a surrogate measure of virus neutralization, where results are reported in terms of % ACE2 displacement. *Panel B:* Same data as panel A, but where ACE2 % displacement activities are plotted longitudinally by participant (PLWH in orange, CTRL in blue). The legend is the same as for Figure 1B.

Multivariable analyses confirmed that, among COVID-19-naive participants, HIV infection was not associated with either WT- or BA-1-specific ACE2 displacement activities at 6 months post-third dose (**Supplementary Table 3**). Rather, having received a mRNA-1273 third dose was the only independent correlate of stronger WT-specific ACE2 displacement activity at this time (p=0.0025). There was also no evidence that a low recent or nadir CD4+ T-cell count was associated with lower WT- or BA.1-specific ACE2 displacement activities 6 months post-third dose in COVID-19-naive PLWH (in fact we observed a weak *inverse* relationship between nadir CD4+ T-cell count and Omicron BA.1-specific ACE2 displacement at this time; **Supplementary Figure 1**). This was consistent with prior observations at one month post-third dose, which we attributed to the observation that PLWH with low nadir CD4+ T-cell counts were eligible for the higher (100mcg) third mRNA-1273 dose [14].

Similar to IgG concentrations, a SARS-CoV-2 breakthrough infection markedly boosted WT- and BA.1-specific ACE2 displacement activities at both three and six months post-third dose (all p<0.0001), where activities at six months in this group were overall significantly greater than peak responses induced by three-dose vaccination (both p<0.0001) (**Figure 2A**). For instance, BA.1-specific ACE2 displacement activity was 95.5% (IQR 90.6-97.6) in the hybrid group six months post-third dose, which was 31% higher than that elicited by vaccination alone. Importantly, hybrid response magnitude was comparable between PLWH and controls (all p≥0.2; **Figure 2B)**, though it is again notable that a minority of PLWH and controls did not show appreciable increases in this activity following infection (**Figure 2B**).

### Longitudinal viral neutralization activity following three-dose vaccination

One month post-third dose, WT-specific neutralization activity in PLWH (median reciprocal plasma dilution 320; IQR 160-1280) was slightly higher than in controls (median 320, IQR 160-320; p=0.004) (**Figure 3A**), though this did not remain significant after multivariable adjustment as previously reported [14].. COVID-19-naive PLWH continued to maintain higher WT-specific neutralization throughout follow-up: by three months, neutralization declined to 160 (IQR 80-320) in PLWH compared to 80 (IQR 40-160) in controls (p=0.02), while by six months, neutralization had declined to a median 80 (IQR 35-160) in PLWH and 40 (IQR 20-80) in controls (p=0.006), values that were below the levels originally elicited by two-dose vaccination (both groups p≤0.01). After multivariable adjustment, HIV infection did not remain significantly associated with WT-specific neutralization activity at six months post-second dose; rather a higher number of health conditions was the only variable independently associated with poorer WT-specific neutralization at this time (**Supplemental Table 4**).

**Figure 3.**
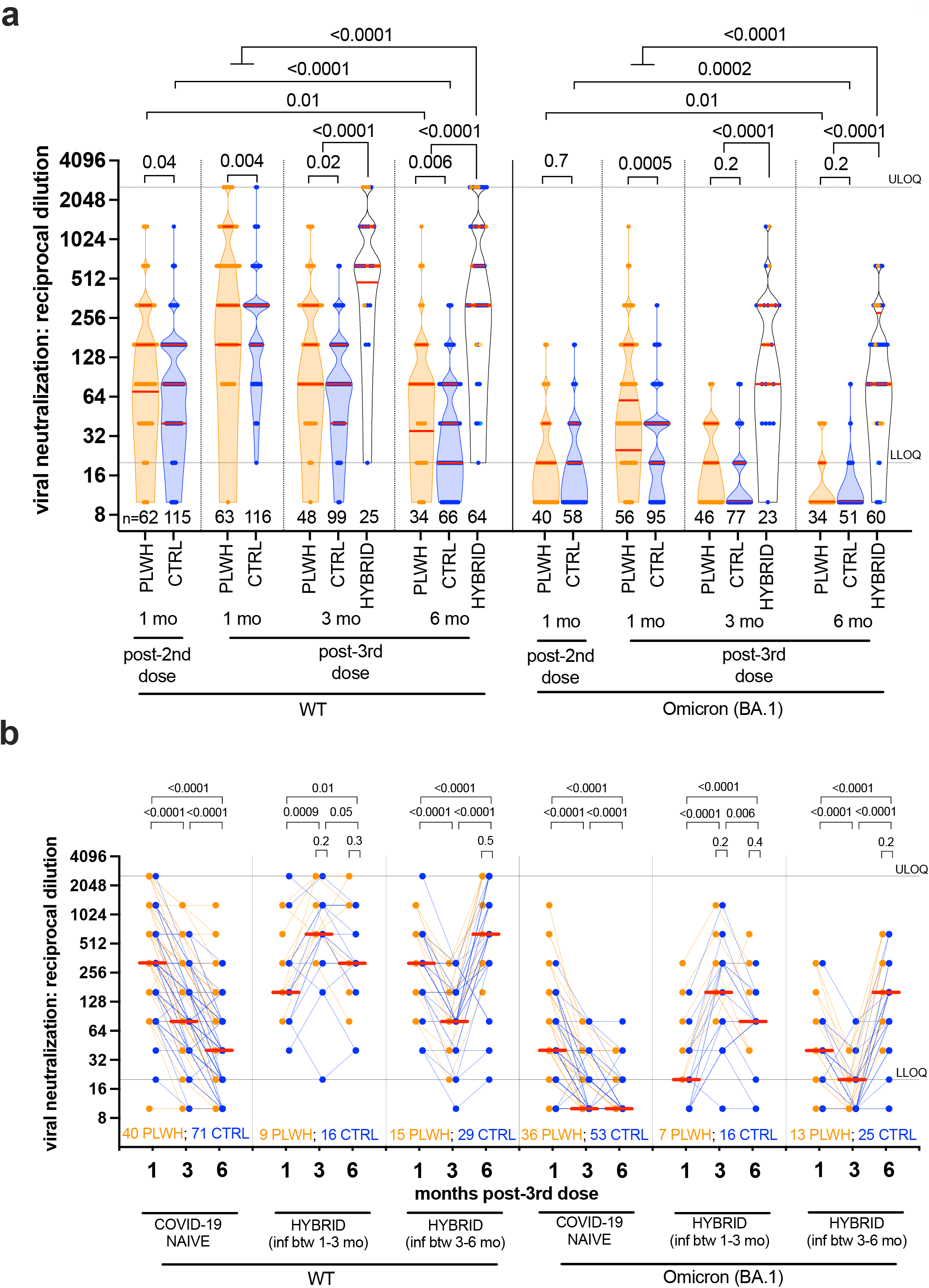
WT- and Omicron BA.1-specific live virus neutralization activity following three-dose COVID-19 vaccination. *Panel A:* Same as Figure 1A, but for live virus neutralization activity, defined as the lowest reciprocal plasma dilution at which neutralization was observed in all wells of a triplicate assay. Serial two-fold dilutions of 1/20 (lower limit of quantification; LLOQ) to 1/2560 (upper limit of quantification; ULOQ) were tested. Plasma samples showing neutralization in fewer than three wells at a 1/20 dilution are displayed as a reciprocal dilution of “10” and were reported as below limit of quantification (BLOQ) in the text. Omicron BA.1 specific neutralization was performed on a subset of samples only. *Panel B:* Same data as panel A, but where neutralization activities are plotted longitudinally by participant (PLWH in orange, CTRL in blue). The legend is the same as for Figure 1B. Note that many data points are superimposed in both panels.

BA.1-specific neutralization was significantly lower than WT responses at all timepoints for all groups (all p<0.0001; not shown), though responses in PLWH were not further impaired compared to controls. In fact, one month post-third dose, BA.1-specific neutralization was higher in PLWH compared to controls: a median 60 (IQR 25 - 160) and 40 (IQR 20 - 40) respectively (p=0.0005), though these values were 5-to-8-fold lower than corresponding WT-specific responses. BA.1-specific neutralization subsequently declined rapidly; at three months, this was BLOQ in 67.4% of COVID-19-naive PLWH and 80.5% of controls (p=0.2), while at six months, this was BLOQ in 82.4% of PLWH and 92.2% of controls (p=0.2), levels that were significantly lower than after two-dose vaccination (all comparisons p≤0.01). In multivariable analyses, older age - but not HIV infection - was the only significant correlate of poorer BA.1-specific neutralization among COVID-19-naive individuals at six month post-third dose (p=0.045; **Supplementary Table 4**). We also observed no significant associations between CD4+ T-cell parameters and either WT- or BA-1-specific neutralization at six months among COVID-19-naive PLWH (p≥0.3; **Supplementary Figure 1**).

By contrast, individuals who experienced breakthrough infection showed significantly stronger WT- and BA.1-specific neutralization compared to their COVID-19-naive counterparts at three and six months (all p<0.0001; **Figure 3A**), responses that were significantly higher than those induced by vaccination alone (all comparisons p≤0.0001) (**Figure 3A**). At six months for example, BA.1-specific responses in participants with hybrid immunity were a median 80 (IQR 80-280), compared to the cohort median of 40 (IQR 20-80) one month post-third dose. Importantly, the magnitude of hybrid immunity was comparable between PLWH and controls at all post-infection time points (all p≥0.2; **Figure 3B**).

### Responses to newer Omicron variants – BA.5

Since the emergence of Omicron BA.1, even more immune evasive variants have developed, including Omicron BA.5 that has dominated globally [22-25]. We therefore longitudinally assessed BA.5-specific neutralization activity in a subset of 18 PLWH and 28 controls who experienced breakthrough infections, which were likely caused by BA.1 or BA.2 based on local epidemiology [17]. One month post-third dose, and prior to SARS-CoV-2 infection, the median reciprocal dilution required for BA.5 neutralization was 20 (IQR BLOQ-20) in this subset, which was two-fold lower than that for BA.1 (median 40, IQR 20-80; p=0.0005), and 160-fold lower than that for WT (median 320, IQR 160-640; p<0.0001) (**Figure 4A**). While neutralization activity against all three virus strains rose significantly post-infection, (all p<0.0001; **Figure 4B**), BA.5-specific neutralization (median 160, IQR 80-200) remained lower than that against BA.1 (median 160, IQR 80-320; p=0.007) and WT (median 640, IQR 320-2560; p<0.0001) (**Figure 4A**). No significant differences were observed between PLWH and controls in their ability to neutralize any virus strains after infection (all p≥0.4) (**Figure 4B**).

**Figure 4.**
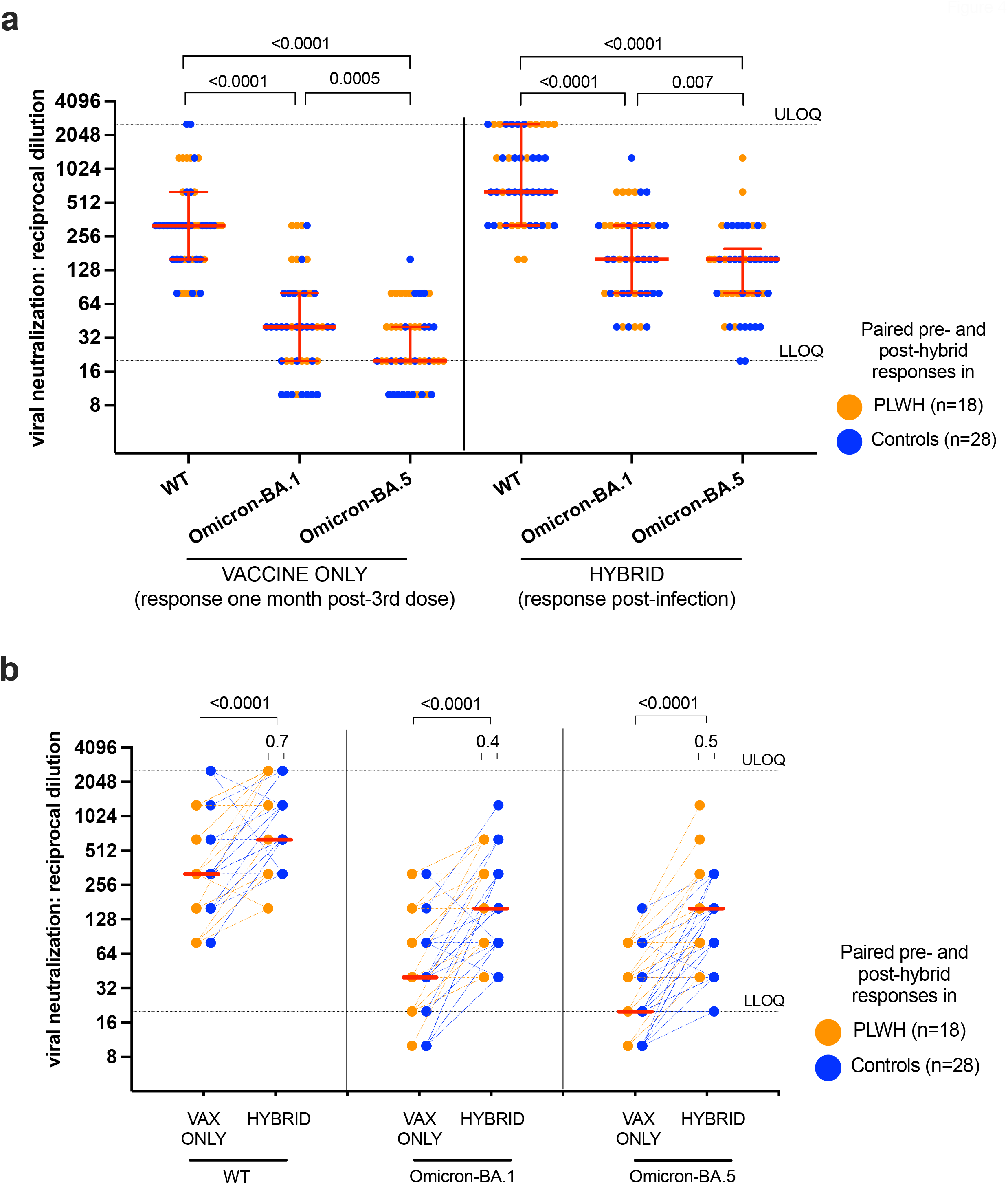
WT-, Omicron BA.1- and Omicron BA.5-specific live virus neutralization activity before and after breakthrough infection. *Panel A:* WT-, Omicron BA.1- and Omicron BA.5-specific neutralization activities in 18 PLWH (orange circles) and 28 CTRL (blue circles) one month after the 3rd vaccine dose (“vaccine only”) and after subsequent SARS-CoV-2 infection (“hybrid”). In this panel, data are plotted to facilitate comparisons between variant-specific neutralization activities, before and after breakthrough infection. P-values, computed on PLWH and CTRL as a combined group, were computed using the Wilcoxon matched pairs test. P-values are not corrected for multiple comparisons. ULOQ/LLOQ: upper/lower limit of quantification. *Panel B*: Same data as in panel A, but plotted longitudinally by participant to highlight the impact of breakthrough infection on neutralization activity. Red lines indicate medians of PLWH and CTRL as a combined group. P-values on top of large brackets compare paired responses before and after breakthrough infection using the Wilcoxon matched pairs test. P-values above small brackets compare responses between PLWH and CTRL at the post-infection time point using the Mann-Whitney U test. Note that a large number of points are superimposed in this plot.

## DISCUSSION

Our results demonstrate that antibody response durability following three-dose COVID-19 vaccination in COVID-19-naive PLWH receiving suppressive ART is comparable to controls without HIV. As we reported previously [14, 16], initial peak antibody responses after three-dose vaccination were similar between PLWH and controls, and our new data additionally show that WT- and Omicron-specific IgG concentrations, ACE2 displacement and virus neutralization activities declined at similar rates among PLWH and controls who remained COVID-19-naive throughout follow-up. Multivariable analyses adjusting for sociodemographic, health and vaccine-related variables confirmed that there was no significant impact of HIV infection on any antibody outcome measure. Nevertheless, by six months post-third dose, all antibody responses in COVID-19-naive participants, regardless of HIV status, had declined to similar or lower levels than those observed at one month after two vaccine doses. In fact, as early as three months post-third dose, BA.1-specific neutralization was already below the limit of quantification in ∼75% of COVID-19-naive participants, regardless of HIV status. Moreover, and consistent with recent reports [26-28], the ability to neutralize BA.5 was even poorer than BA.1.

By contrast, almost all participants who experienced their first SARS-CoV-2 infection (presumably BA.1 or BA.2 [17]) post-third dose displayed enhanced antibody responses to all viral variants tested. In fact, at 6 months post-third dose, all antibody measures in breakthrough infection cases were on average higher than those elicited by vaccination alone. Importantly, the magnitude of hybrid responses in PLWH was consistently comparable to those of controls. Nevertheless, it was notable that responses to BA.5 remained significantly lower than those against WT and BA.1 even after infection, and that a minority of participants failed to show improved antibody responses post-infection, a phenomenon which requires further study.

Our study has several limitations. Our observations may not be generalizable to PLWH with low CD4+ T-cell counts and/or who are not receiving suppressive ART. Indeed, results from several studies indicate that PLWH with CD4+ T-cell counts below 500 cells/mm^3^ mount weaker responses to the first [29-31] and second [30-37] doses of COVID-19 vaccine. In addition, we did not assess cellular immunity in participants. Furthermore, any recommendations based on our results are limited by the emergence of new SARS-CoV-2 variants, including strains that may be more immune-evasive than BA.5 (*e*.*g*. BA.2.75) [38-42].

## CONCLUSIONS

Our observations confirm the humoral immune benefits of third COVID-19 vaccine doses in PLWH receiving suppressive ART, and further reveal that the durability of third-dose responses is comparable to that in persons without HIV. Nevertheless, regardless of HIV status, individuals who remain COVID-19-naive will benefit from a fourth dose within 3-6 months of their third dose, since antibody concentrations and neutralization activities declined markedly over time in this group, and the ability of vaccine-induced responses to neutralize the dominant Omicron BA.5 variant were even poorer than BA.1. By contrast, the majority of individuals who experienced their first SARS-CoV-2 infection post-third vaccine dose showed significantly higher antibody activities than those induced by vaccination alone, though anti-BA.5 responses remained weaker than anti-BA.1, even after breakthrough infection. Nevertheless, these observations suggest that a slightly delayed fourth dose (*e*.*g*. to 3-6 months following infection) would optimally benefit this group. Further studies of hybrid response durability are required, as are direct comparisons with immune responses elicited by a fourth dose in COVID-19-naїve individuals, particularly in light of new bivalent formulations that include WT and Omicron Spike antigens.

## Data Availability

All data produced in the present study are available upon reasonable request to the authors.

## ACKNOWLEDGEMENTS

This work is dedicated to the memory of our friend and colleague Hesham Ali who sadly passed away in July 2022. We thank the phlebotomists and laboratory staff at the BC Centre for Excellence in HIV/AIDS, the Hope to Health Research and Innovation Centre, St. Paul’s Hospital, and Simon Fraser University for assistance. Above all, we thank the participants, without whom this study would not have been possible.

## Supplemental Figures

**Supplemental Figure 1**: Relationships between most recent and nadir CD4+ T-cell counts and humoral responses six months post-third vaccine dose. Relationships were assessed using Spearman’s correlation. Measurements against wild-type (WT) SARS-CoV-2 are indicated by black symbols; measurements against Omicron BA.1 are indicated by open symbols. Analyses are restricted to COVID-19-naїve PLWH. ULOQ/LLOQ: upper/lower limit of quantification

**Supplemental Figure 2: Temporal declines in WT- and Omicron BA.1-specific anti-RBD IgG concentrations following three-dose COVID-19 vaccination**. *Panel A:* Temporal declines in WT-specific anti-RBD IgG concentrations following three-dose vaccination in PLWH (orange) and CTRL (blue) who remained COVID-19-naїve during follow-up, and who completed all post-third dose visits. *Panel B*: Estimated WT-specific IgG half-lives following three-dose vaccination, calculated by fitting an exponential curve to each participant’s data in panel B. Red bars and whiskers represent the median and IQR. P-values were computed using the Mann-Whitney U-test. *Panel C*: Same as panel A, but for Omicron (BA.1)-specific anti-RBD IgG concentration. *Panel D*: Same as panel B, but for half-lives calculated from the BA.1-specific data.

