## Supplemental Tables for "Antibody response durability following three-dose COVID-19 vaccination in people with HIV receiving suppressive ART"

**Supplementary Table 1: Multivariable linear regression analysis of the relationship between sociodemographic, health and vaccine-related variables on WT- and BA.1-specific IgG concentrations six months post-third COVID-19 vaccine dose.**

| **Immunogenicity outcome**^a^ | **Variable** | **SARS-CoV-2 Variant** | | | | | |
| --- | --- | --- | --- | --- | --- | --- | --- |
|  |  | **Wild-type** | | | **Omicron BA.1** | | |
|  |  | **Estimate** | **95% CI** | **p-value** | **Estimate** | **95% CI** | **p-value** |
| **Anti-RBD IgG (log_10_BAU/ml)** | HIV infection | 0.14 | -0.12 to 0.39 | 0.29 | 0.23 | -0.019 to 0.49 | 0.069 |
| **6 months post-third dose** | Age (per year) | 0.0021 | -0.0042 to 0.0084 | 0.5 | 0.0018 | -0.0044 to 0.0081 | 0.56 |
|  | Male sex | -0.21 | -0.43 to 0.0083 | 0.059 | -0.28 | -0.5 to -0.064 | **0.012** |
|  | White ethnicity | -0.092 | -0.29 to 0.1 | 0.35 | 0.0022 | -0.19 to 0.2 | 0.98 |
|  | # Chronic conditions (per additional) | -0.14 | -0.26 to -0.015 | **0.028** | -0.15 | -0.27 to -0.029 | **0.016** |
|  | Dual ChAdOx1 as initial regimen | -0.089 | -0.42 to 0.24 | 0.59 | -0.16 | -0.49 to 0.17 | 0.33 |
|  | mRNA-1273 as 3rd dose | 0.22 | 0.023 to 0.41 | **0.029** | 0.15 | -0.044 to 0.34 | 0.13 |
|  | Interval btw 2nd and 3rd doses (per day) | 0.0011 | -0.0018 to 0.004 | 0.44 | 0.0015 | -0.0014 to 0.0044 | 0.3 |

^a^Analysis was restricted to participants who remained COVID-19 naïve six months post-third vaccine dose

**Supplementary Table 2: Multivariable linear regression analysis of the relationship between sociodemographic, health and vaccine-related variables on WT- and BA.1-specific IgG half-lives following three-dose COVID-19 vaccination.**

| **Immunogenicity outcome**^a^ | **Variable** | **SARS-CoV-2 Variant** | | | | | |
| --- | --- | --- | --- | --- | --- | --- | --- |
|  |  | **Wild-type** | | | **Omicron BA.1** | | |
|  |  | **Estimate** | **95% CI** | **p-value** | **Estimate** | **95% CI** | **p-value** |
| **Anti-RBD IgG half-lives** | HIV infection | 1.7 | -25 to 28 | 0.9 | 12 | -18 to 42 | 0.42 |
| **post-3rd dose, in days** | Age (per year) | 0.41 | -0.26 to 1.1 | 0.23 | 0.33 | -0.42 to 1.1 | 0.38 |
|  | Male sex | -21 | -44 to 2.1 | 0.075 | -12 | -39 to 14 | 0.35 |
|  | White ethnicity | 3.9 | -17 to 25 | 0.71 | 6.5 | -17 to 30 | 0.58 |
|  | # Chronic conditions (per additional) | -8.3 | -21 to 4.3 | 0.19 | -9.1 | -23 to 5.1 | 0.21 |
|  | Dual ChAdOx1 as initial regimen | -0.31 | -35 to 35 | 0.99 | -2.7 | -42 to 37 | 0.89 |
|  | mRNA-1273 as 3rd dose | 14 | -6.7 to 34 | 0.18 | -3.9 | -27 to 19 | 0.74 |
|  | Interval btw 2nd and 3rd doses (per day) | 0.011 | -0.3 to 0.32 | 0.94 | -0.066 | -0.41 to 0.28 | 0.7 |

^a^Analysis was restricted to participants who remained COVID-19 naïve throughout follow-up, and who completed all three post-third dose study visits

**Supplementary Table 3: Multivariable linear regression analysis of the relationship between sociodemographic, health and vaccine-related variables on WT- and BA.1-specific ACE2 displacement activity six months post-third COVID-19 vaccine dose.**

| **Immunogenicity outcome** | **Variable** | **SARS-CoV-2 Variant** | | | | | |
| --- | --- | --- | --- | --- | --- | --- | --- |
|  |  | **Wild-type** | | | **Omicron BA.1** | | |
|  |  | **Estimate** | **95% CI** | **p-value** | **Estimate** | **95% CI** | **p-value** |
| **ACE2 Displacement (%)** | HIV infection | -1.2 | -11 to 9.1 | 0.82 | 11 | -2.6 to 25 | 0.11 |
|  | Age (per year) | -0.068 | -0.32 to 0.19 | 0.6 | -0.23 | -0.57 to 0.11 | 0.19 |
|  | Male sex | -4.3 | -13 to 4.7 | 0.34 | -11 | -23 to 1 | 0.072 |
|  | White ethnicity | 0.68 | -7.2 to 8.6 | 0.86 | -1.1 | -12 to 9.4 | 0.84 |
|  | # Chronic conditions (per additional) | -4.2 | -9 to 0.73 | 0.094 | -4.2 | -11 to 2.3 | 0.2 |
|  | Dual ChAdOx1 as initial regimen | -3.5 | -17 to 10 | 0.61 | 0.53 | -17 to 18 | 0.95 |
|  | mRNA-1273 as 3rd dose | 12 | 4.5 to 20 | **0.0025** | 5 | -5.6 to 16 | 0.35 |
|  | Interval btw 2nd and 3rd doses (per day) | 0.056 | -0.061 to 0.17 | 0.35 | -0.0051 | -0.16 to 0.15 | 0.95 |

^a^Analysis was restricted to participants who remained COVID-19 naïve six months post-third vaccine dose

**Supplemental Table 4: Multivariable analyses of the relationship between sociodemographic, health and vaccine-related variables on viral neutralization six months following the third vaccine dose.**

| **Immunogenicity outcome^a^** | **Variable** | **SARS-CoV-2 Variant** | | | | | |
| --- | --- | --- | --- | --- | --- | --- | --- |
|  |  | **Wild-type^c^** | | | **Omicron BA.1^c^** | | |
|  |  | **Estimate ^c^** | **95% CI ^c^** | **p-value** | **Odds Ratio ^c^** | **95% CI^c^** | **p-value** |
| **Log_2_ viral neutralization^b^** | HIV infection | 0.96 | -0.36 to 2.3 | 0.15 | 4.9 | 0.67 to 51 | 0.14 |
|  | Age (per year) | -0.0014 | -0.026 to 0.023 | 0.91 | 0.96 | 0.91 to 1 | **0.045** |
|  | Male sex | -0.52 | -1.3 to 0.27 | 0.2 | 0.28 | 0.052 to 1.2 | 0.1 |
|  | White ethnicity | -0.54 | -1.2 to 0.14 | 0.12 | 1.3 | 0.39 to 4.3 | 0.69 |
|  | # Chronic conditions (per additional) | -0.46 | -0.88 to -0.047 | **0.029** | 0.77 | 0.31 to 1.8 | 0.55 |
|  | Dual ChAdOx1 as initial regimen | 0.72 | -0.46 to 1.9 | 0.23 | 3 | 0.52 to 18 | 0.21 |
|  | mRNA-1273 as 3rd dose | 0.36 | -0.32 to 1 | 0.29 | 0.93 | 0.28 to 3.3 | 0.91 |
|  | Interval btw 2nd and 3rd doses (per day) | -0.0019 | -0.012 to 0.0084 | 0.71 | 0.99 | 0.98 to 1 | 0.55 |
|  | EDTA as anticoagulant^d^ | 0.27 | -1.2 to 1.7 | 0.71 | 0.7 | 0.057 to 7.1 | 0.77 |

^a^Analysis was restricted to participants who remained COVID-19 naïve six months post-third vaccine dose

^b^reciprocal plasma dilutions were log_2_ transformed prior to multivariable analysis.

^c^Multivariable analysis for wild-type neutralization was performed using linear regression. Multivariable analysis for Omicron BA.1 neutralization was performed using logistic regression due to a high proportion of measurements below the limit of quantification (BLOQ) at this time point.

^d^Neutralization assays were performed using plasma (not serum as in other analyses), so the models additionally correct for the anticoagulant.
