## Supplementary figures and images for "Antibody response durability following three-dose COVID-19 vaccination in people with HIV receiving suppressive ART"

### Supplemental Figure S1

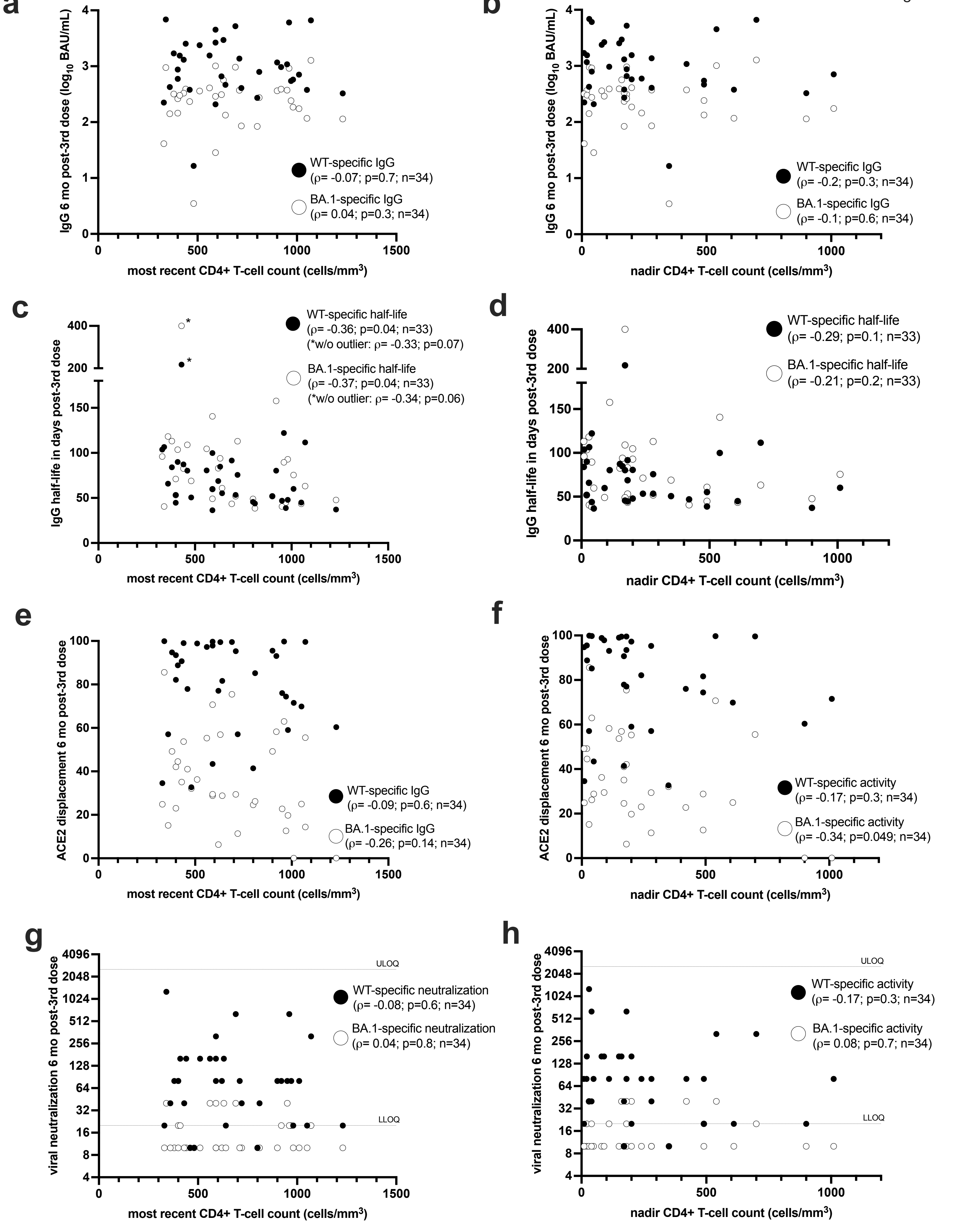

### Supplemental Figure S2

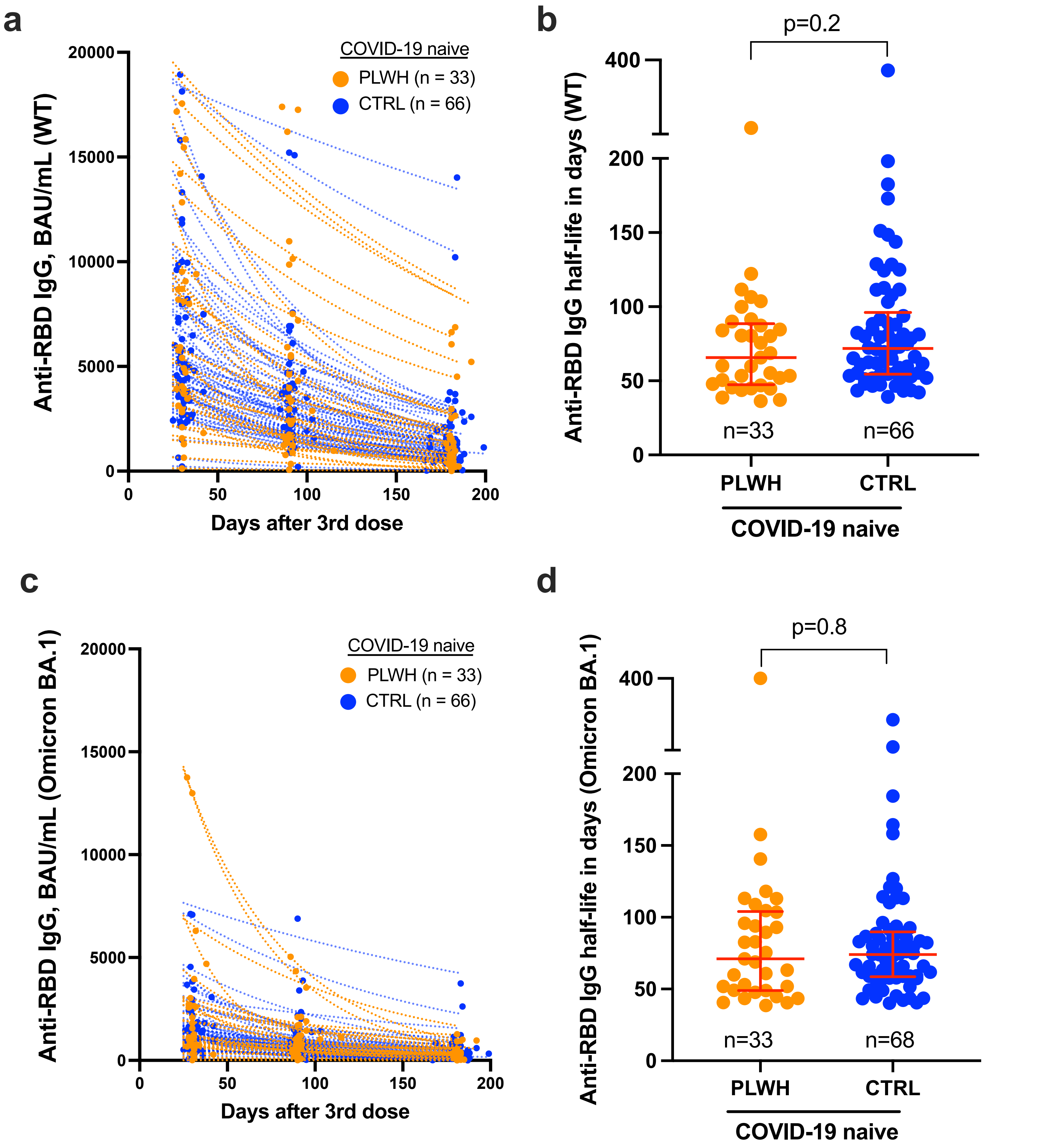
